# Influenza Vaccine Effectiveness Against Influenza A(H3N2)-Related Illness in the United States During the 2021–2022 Influenza Season

**DOI:** 10.1101/2022.10.05.22280702

**Authors:** Ashley M. Price, Brendan Flannery, H. Keipp Talbot, Carlos G. Grijalva, Karen J. Wernli, C. Hallie Phillips, Arnold S. Monto, Emily T. Martin, Edward A. Belongia, Huong Q. McLean, Manjusha Gaglani, Manohar Mutnal, Krissy Moehling Geffel, Mary Patricia Nowalk, Sara Y. Tartof, Ana Florea, Callie McLean, Sara S. Kim, Manish M. Patel, Jessie R. Chung

## Abstract

**Background:** In the United States, influenza activity during the 2021-2022 season was modest and sufficient enough to estimate influenza vaccine effectiveness for the first time since the beginning of the COVID-19 pandemic. We estimated influenza vaccine effectiveness against lab-confirmed outpatient acute illness caused by predominant A(H3N2) viruses.

**Methods:** Between October 2021 and April 2022, research staff across 7 sites enrolled patients aged ≥6 months seeking outpatient care for acute respiratory illness with cough. Using a test-negative design, we assessed VE against influenza A(H3N2). Due to strong correlation between influenza and SARS-CoV-2 vaccination, participants who tested positive for SARS-CoV-2 were excluded from vaccine effectiveness estimations. Estimates were adjusted for site, age, month of illness, race/ethnicity and general health status.

**Results:** Among 6,260 participants, 468 (7%) tested positive for influenza only, including 440 (94%) for A(H3N2). All 206 sequenced A(H3N2) viruses were characterized as belonging to genetic group 3C.2a1b subclade 2a.2, which has antigenic differences from the 2021–2022 season A(H3N2) vaccine component that belongs to clade 3C.2a1b subclade 2a.1. After excluding 1,948 SARS-CoV-2 positive patients, 4,312 patients were included in analyses of influenza VE; 2,463 (57%) were vaccinated against influenza. Effectiveness against A(H3N2) for all ages was 36% (95%CI, 20-49%) overall; 40% (95%CI, 24-53%) for those aged 6 months-49 years; and 10% (95%CI, -60-49%) for those aged ≥50 years.

**Conclusion:** Influenza vaccination in 2021–2022 provided protection against influenza A(H3N2)-related outpatient visits among young persons, with no measurable protection among older adults.

## Introduction

Global influenza activity declined to historically low levels after the beginning of the SARS-CoV-2 pandemic in March 2020. In the United States (US), influenza activity increased in late 2021, with two distinct periods of influenza virus circulation. The first period began November 2021 and continued through mid-January 2022. Influenza activity declined from late December 2021 through late January 2022 during the rapid rise in the B.1.1.529 (Omicron) SARS-CoV-2 variant [1, 2]. In the second period, starting mid-January, influenza activity continued at a low level through June 2021, with a second peak from mid-March to mid-May which was much later than previous influenza seasons. Despite the low to moderate level of influenza activity throughout the season, there were an estimated 8.0–13.0 million influenza illnesses, 3.7–6.1 million influenza-related medical visits, 82,000–170,000 hospitalizations, and 5,000–14,000 influenza-associated deaths during 2021–2022 [3].

Annual influenza vaccination starting at 6 months of age is recommended in the US as the most effective means of mitigating and preventing influenza-associated illnesses and complications. Influenza vaccination is especially important during the ongoing COVID-19 pandemic to reduce the burden of medical visits and hospitalizations due to respiratory illness [4]. However, in November 2021, early influenza activity suggested potentially reduced vaccine effectiveness (VE) against predominant A(H3N2) viruses belonging to the 3C.2a1b.2a.2 subclade [1, 5]. Low global influenza circulation during the 2020– 2021 influenza season limited selection of vaccine reference viruses for the 2021–2022 Northern Hemisphere influenza season [6]. Since 2004–2005, CDC and collaborating study sites have produced annual estimates of influenza vaccine effectiveness against laboratory-confirmed, mild to moderate (outpatient) medically attended acute respiratory infection (ARI) for every season except 2020–2021 due to very low influenza activity. Here, we report VE estimates for the 2021–2022 season against A(H3N2)-associated influenza illness, including VE estimates by age group and early-versus late-season influenza activity.

## Methods

### Study Population

This study was conducted within the US Flu VE Network, which consists of participating health systems in 7 states: California, Michigan, Pennsylvania, Tennessee, Texas, Washington, and Wisconsin. Details of this network have been described previously [1, 7]. Between October 4, 2021, and April 30, 2022, research staff screened patients aged ≥6 months who had ARI with cough, fever/feverishness, or loss of taste or smell seeking outpatient medical care (i.e., telehealth, primary care, urgent care, or emergency department) or clinical testing for SARS-CoV-2 ≤10 days after illness onset [8]. Research staff interviewed participants using standard questionnaires for data on patient demographics, symptoms, subjective general health status before illness onset, and self-or parent-reported receipt of the 2021– 2022 seasonal influenza vaccine.

At enrollment, study staff collected nasal and/or oropharyngeal swab specimens (only nasal swab specimens were collected for children aged <2 years). Specimens were tested for influenza and SARS-CoV-2 using real-time reverse-transcriptase polymerase chain reaction (RT-PCR). Influenza-positive specimens with RT-PCR cycle threshold values <30 were sent to CDC for whole genome sequencing [9]. Data sources for documentation of influenza and COVID-19 vaccination included electronic medical records and immunization information systems. Self-reported vaccination was considered plausible evidence of influenza vaccine receipt for participants who specified location and timing of influenza vaccine receipt. This activity was reviewed and approved by the CDC and each US Flu VE Network site’s Institutional Review Board.

### Vaccine Effectiveness Estimates

Vaccine effectiveness (VE) was estimated from logistic regression models using the test-negative design as 100% x (1 – adjusted odds ratio [OR]) [10]. We included participants who reported cough or fever/feverishness and were tested within 7 days of illness onset. Cases were patients testing positive for influenza by RT-PCR, and controls were patients testing negative for both influenza and SARS-CoV-2. Due to strong correlation between influenza and SARS-CoV-2 vaccination, participants who tested RT-PCR positive for SARS-CoV-2 infection were excluded from influenza VE estimation [11]. We conducted a sensitivity analysis including participants who tested positive for SARS-CoV-2.

Influenza vaccination status was determined from documentation from electronic sources or from self-report of vaccination with a date of administration. receipt of one or more doses of any 2021– 2022 seasonal influenza vaccine ≥14 days prior to illness onset. Using multivariable logistic regression, odds ratio estimates were adjusted for study site, age, month of illness onset, race/ethnicity, and general health status. A 95% confidence interval (CI) was calculated for each estimate. Stratified analyses were performed by age group where sample size permitted [12]. We also separately examined VE during October 4, 2021—January 15, 2022, corresponding to the first period of seasonal influenza activity until widespread circulation of the SARS-CoV-2 Omicron variant, and January 16—April 30, 2022, during the second, extended period of influenza virus circulation. Analyses were conducted using SAS 9.4 software (SAS Institute) and R software, version 4.0.2 (R Foundation for Statistical Computing).

## Results

Among 8917 participants with ARI enrolled at the seven study sites during October 4, 2021–April 30, 2022, 2657 (30%) were excluded due to enrollment outside periods of influenza circulation for each site (n=1133), influenza vaccination <14 days before illness onset (n=143), missing testing or vaccination data (n=628), not meeting the clinical ARI definition (n=753), or co-detection of influenza and SARS-CoV-2 (n=16). Weekly influenza positivity ranged from 0% to <10% throughout the period; SARS-CoV-2 positivity ranged from 10% to 60%, with a peak in early to mid-January 2022 when the SARS-CoV-2 Omicron variant predominated (Figure 1). Among 6260 participants, 468 (7%) tested positive for influenza, 1948 (31%) tested positive for SARS-CoV-2, and 3844 (62%) tested negative for both influenza and SARS-CoV-2. Among influenza-positive participants, 440 (94%) were subtyped as A(H3N2), 2 (<1%) were A(H1N1)pdm09, and 26 (6%) were influenza A with no subtype result. No influenza B cases were detected. A total of 206 (47%) A(H3N2) viruses were characterized by whole genome sequencing; all belonged to genetic group 3C.2a1b subclade 2a.2 (full clade: 3C.2a1b.2a.2). The median age of influenza-positive cases (19 years) was younger than the median age of SARS-CoV-2 positive participants (37 years) or test-negative controls (33 years) (Table 1). The proportion of patients with influenza differed by study site, sex, age group, race/ethnicity, days from illness onset to enrollment, general health status, and presence of high-risk medical conditions. (Table 1).

**TABLE 1.**
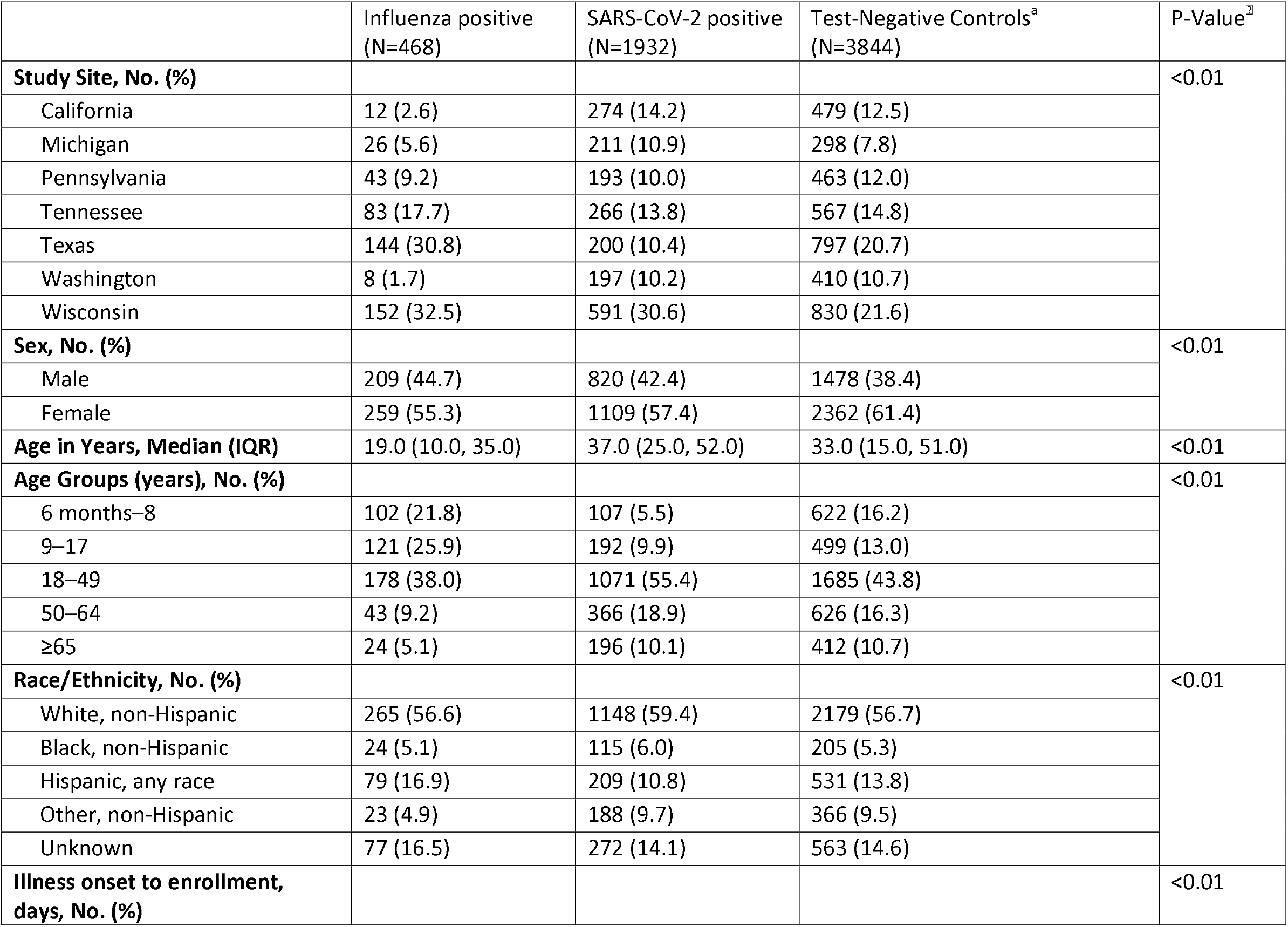
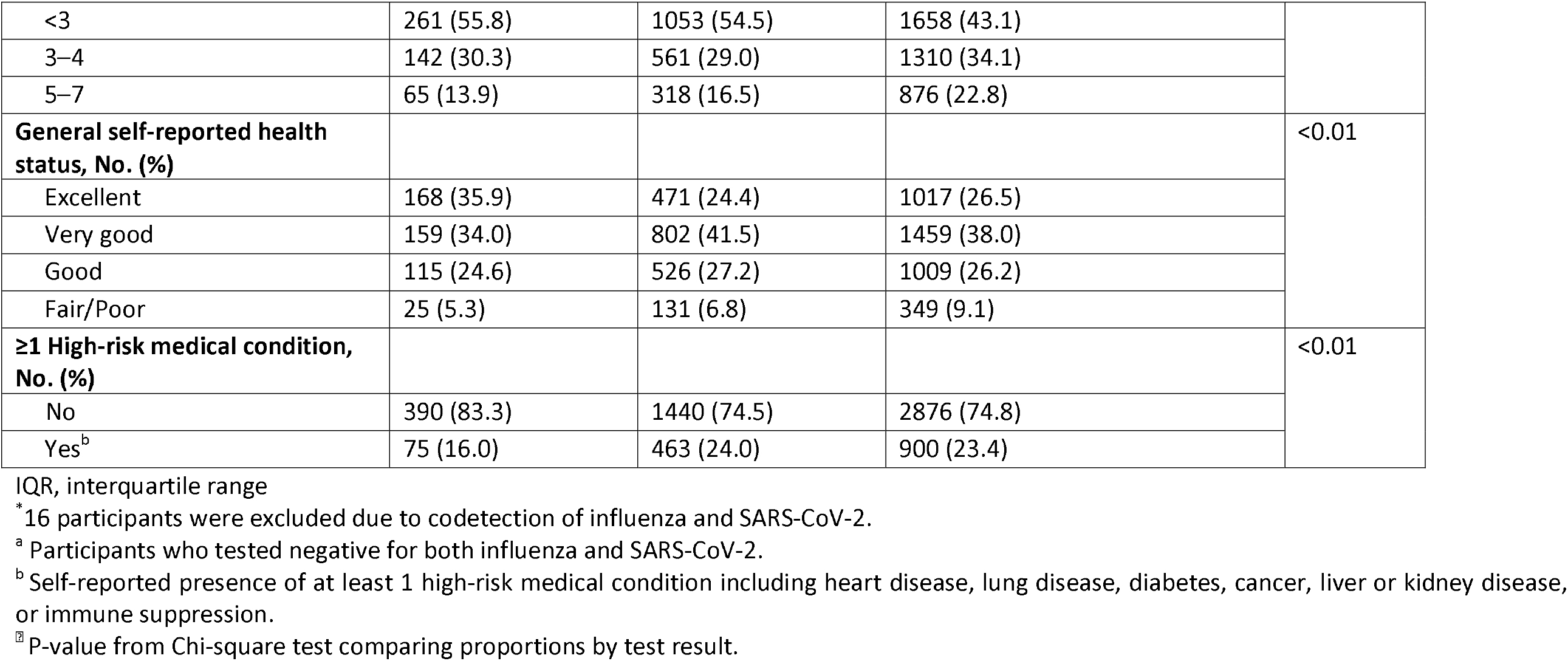
Characteristics of participants with medically attended acute respiratory infection by influenza and SARS-CoV-2 test result, October 4, 2021 – April 30, 2022^*^

**FIGURE 1.**
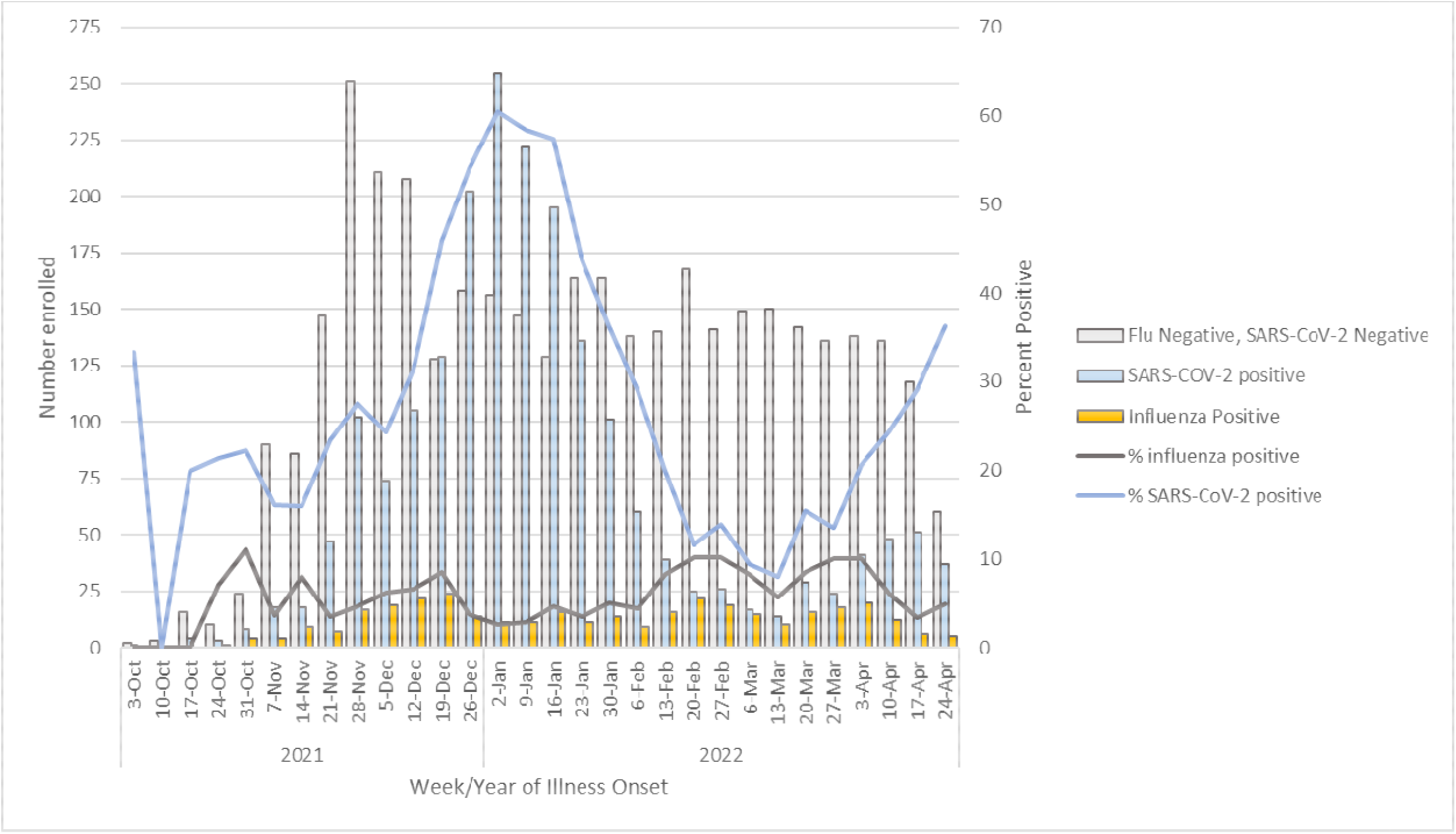
Distribution of participants by test result and percent positivity by week of onset, October 4, 2021 – April 30, 2022

After excluding SARS-CoV-2 positive patients who were tested for influenza, 4312 patients were included in analyses of influenza VE; 2463 (57%) were vaccinated against influenza (Table 2). Compared to unvaccinated participants, participants who received influenza vaccine were older, more likely to be non-Hispanic white, more likely to have received 3 or more COVID-19 vaccinations, and more likely to report having at least one high-risk medical condition (Table 2).

**TABLE 2.** Characteristics of participants by influenza vaccination status, October 4, 2021 – April 30, 2022^*^

|  | Vaccinated*<br>(N=2463) | Unvaccinated<br>(N=1849) | P-Value<sup>§</sup> |
| --- | --- | --- | --- |
| **Study Site, No. (%)** |  |  | <0.01 |
| California | 333 (13.5) | 158 (8.5) |  |
| Michigan | 257 (10.4) | 67 (3.6) |  |
| Pennsylvania | 278 (11.3) | 228 (12.3) |  |
| Tennessee | 371 (15.1) | 279 (15.1) |  |
| Texas | 351 (14.3) | 590 (31.9) |  |
| Washington | 290 (11.8) | 128 (6.9) |  |
| Wisconsin | 583 (23.7) | 399 (21.6) |  |
| **Sex, No. (%)** |  |  | <0.01 |
| Male | 919 (37.3) | 768 (41.5) |  |
| Female | 1542 (62.6) | 1079 (58.4) |  |
| **Age in Years, Median (IQR)** | 36.0 (15.0, 56.0) | 25.0 (12.0, 41.0) | <0.01 |
| **Age Groups (years), No. (%)** |  |  | <0.01 |
| 6 months–8 | 394 (16.0) | 330 (17.8) |  |
| 9–17 | 255 (10.4) | 365 (19.7) |  |
| 18–49 | 1010 (41.0) | 853 (46.1) |  |
| 50–44 | 454 (18.4) | 215 (11.6) |  |
| ≥65 | 350 (14.2) | 86 (4.7) |  |
| **Race/Ethnicity, No. (%)** |  |  | <0.01 |
| White, non-Hispanic | 1483 (60.2) | 961 (52.0) |  |
| Black, non-Hispanic | 93 (3.8) | 136 (7.4) |  |
| Hispanic, any race | 278 (11.3) | 332 (18.0) |  |
| Other, non-Hispanic | 243 (9.9) | 146 (7.9) |  |
| Unknown | 366 (14.9) | 274 (14.8) |  |
| **Illness onset to enrollment, days, No. (%)** |  |  | 0.44 |
| <3 | 1112 (45.1) | 807 (43.6) |  |
| 3–4 | 810 (32.9) | 642 (34.7) |  |
| 5–7 | 541 (22.0) | 400 (21.6) |  |
| **General self-reported health status, No. (%)** |  |  | 0.06 |
| Excellent | 655 (26.6) | 530 (28.7) |  |
| Very good | 942 (38.2) | 676 (36.6) |  |
| Good | 637 (25.9) | 487 (26.3) |  |
| Fair/Poor | 221 (9.0) | 153 (8.3) |  |
| **≥1 High-risk medical condition present, No. (%)** |  |  | <0.01 |
| No | 1764 (71.6) | 1502 (81.2) |  |
| Yes<sup>a</sup> | 653 (26.5) | 322 (17.4) |  |
| **COVID-19 Vaccination Status, No. (%)** |  |  |  |
| 0 doses | 211 (8.6) | 666 (36.0) | <0.01 |
| 1–2 doses | 652 (26.5) | 679 (36.7) | <0.01 |
| ≥3 doses | 1299 (52.7) | 268 (14.5) | <0.01 |
| **Influenza rRT-PCR result, No. (%)** |  |  |  |
| Negative | 2265 (92.0) | 1579 (85.4) | <0.01 |
| Influenza A | 198 (8.0) | 270 (14.6) |  |
| A(H1N1)pdm09 | 0 (0) | 2 (0.1) |  |
| A(H3N2) | 182 (7.4) | 258 (14.0) |  |
| Influenza B | 0 (0) | 0 (0) |  |

IQR, interquartile range
<sup>a</sup>Participants who tested positive for SARS-CoV-2 are excluded.
<sup>b</sup> Self-reported presence of at least 1 high-risk medical condition including heart disease, lung disease, diabetes, cancer, liver or kidney disease, or immune suppression.
<sup>‡</sup> P-value from Chi-square test comparing proportions by test result.

For all ages combined, VE against influenza A was 36% (95% CI, 21%–48%) and 36% (95% CI, 20%–49%) for A(H3N2), specifically. VE against A(H3N2) varied by age from 51% (95%CI, 19%–70%) among patients aged 6 months – 8 years, 32% (95%CI, 3%–52%) among adults aged 18–49 years, and 10% (95%CI, -60%–49%) among adults aged ≥50 years (Figure 2). We were underpowered to detect a statistically significant VE of 30% in all age groups [12], but numbers of cases among older adults aged ≥50 years was particularly sparse.

**FIGURE 2.**
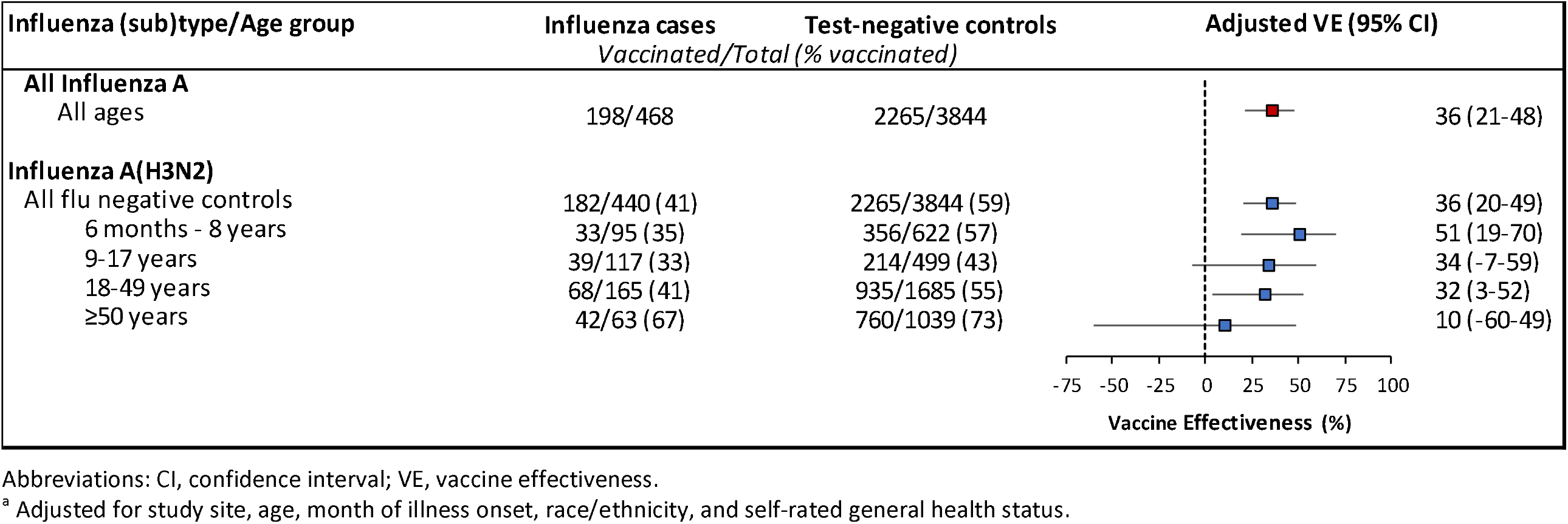
Vaccine effectiveness against outpatient influenza A- and A(H3N2)-associated illness, October 4, 2021 – April 30, 2022

In a sensitivity analysis including participants who tested positive for SARS-CoV-2 at enrollment, VE was slightly lower (30% [95% CI, 14%–43%]). There was no statistically significant difference observed between VE among those enrolled on or before January 15, 2022 (29% [95% CI, –5%–53%]) compared to those enrolled after January 15, 2022 (37% [95% CI, 19%–51%]) (Figure 3).

**FIGURE 3.**
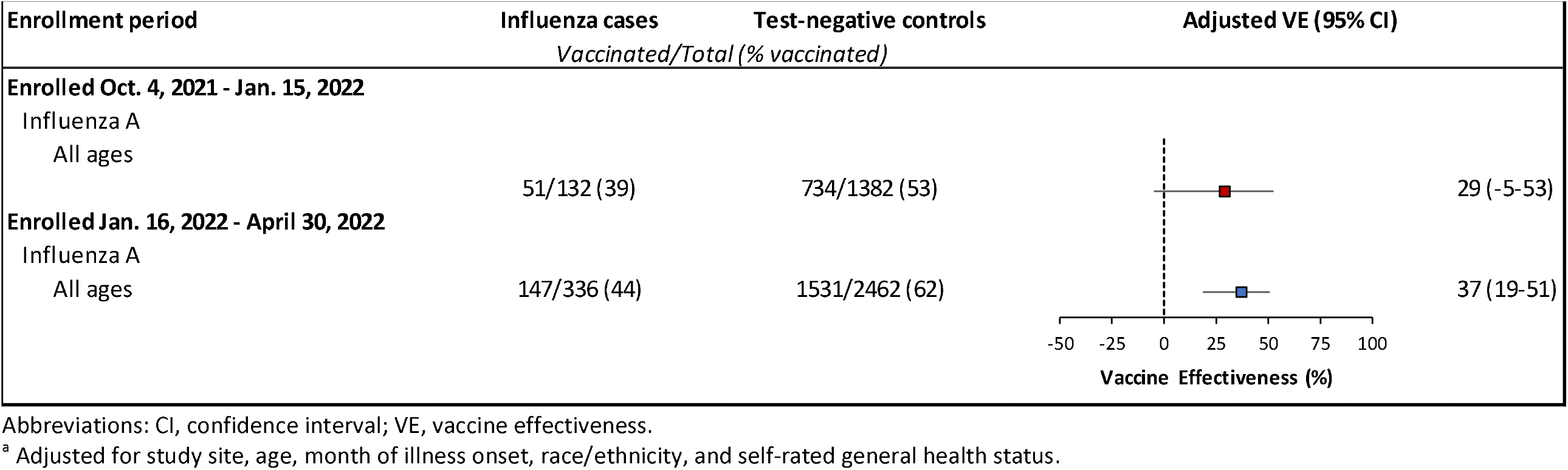
Vaccine effectiveness ^a^ against outpatient influenza A(H3N2)-associated illness by time since vaccination, October 4, 2021 – April 30, 2022, United States

## Discussion

For the 2021–2022 influenza season, influenza A(H3N2) viruses predominated although virus circulation remained relatively low in comparison to the prevalence of SARS-CoV-2 Omicron variant and subvariants (B.1.1.529/BA.1/BA.2). Influenza vaccines were 36% effective against A(H3N2)-related illnesses among all participants less than 50 years of age. However, we were unable to detect statistically significant protection against laboratory-confirmed influenza among adults aged 50 years and older. In general, detection of statistically significant VE below 30% with high vaccine coverage requires a larger sample size than we were able to enroll this season [12].

Lower VE among older adults compared with younger persons has been observed in previous seasons, especially against A(H3N2) viruses [13]. The A(H3N2) reference virus selected for egg-based and cell-or recombinant-based 2021–2022 influenza vaccines, A/Cambodia/e0826360/2020, belonged to clade 3C.2a1b subclade 2a.1, while almost all influenza A(H3N2) viruses circulating throughout the US from November 2021–April 2022 belonged to the 3C.2a1b subclade 2a.2. While these two subclades were genetically similar, there were antigenic differences detected between circulating viruses and the cell- and egg-grown vaccine component, based on post-infection ferret antibody cross-reactivity [6]. It is unclear whether older adults responded differently than younger persons to the mismatched A(H3N2) vaccine component or if there were differences in cross-protective antibodies. Alternatively, influenza-positive older patients may have differed in unmeasured characteristics from those testing negative for influenza and SARS-CoV-2 in ways that affected VE estimates. It is also unclear whether egg-adapted changes in A(H3N2) viruses used in vaccine production or whether boosting antibodies specific to egg-adapted viruses may have affected VE, although statistically significant VE was observed among young children and adults aged 18–49 years. At the time of this report, we were unable to calculate VE by vaccine type because complete vaccination data including vaccine type were not yet available. In recent influenza seasons, uptake of recombinant influenza vaccine and cell-culture influenza vaccine has increased among US Flu VE Network participants including older adults, who also may receive high-dose egg-based influenza vaccine [14].

SARS-CoV-2-positive participants were excluded from estimates of influenza VE to remove potential bias resulting from positive correlation between COVID-19 vaccination and influenza vaccination [11]. A previous study carried out nearly a decade ago has shown that the test-negative design produced similar estimates of influenza VE when test-negative controls included participants who tested positive for other respiratory viruses [15]. However, in that study, vaccines were not available for the other respiratory viruses detected. In our study, SARS-CoV-2-positive participants were less likely to receive influenza vaccine compared to those who tested negative for SARS-CoV-2. This resulted in lower estimated influenza VE when SARS-CoV-2-positive participants were included. The effect of this bias was greatest during the period of high COVID-19 prevalence. Estimated mid-season influenza VE from October 2021–February 2022 was 14% (95%CI, -17%–37%) [1]. After removing SARS-CoV-2-positive influenza negative controls, we estimate the corrected VE for this period was approximately 30%. Over the entire study period October 2021–April 2022, removal of SARS-CoV-2-positive participants increased VE point estimates >5% but <10%.

Several limitations of our study should be considered. First, the validity of observational VE studies depends on accurate classification of vaccination status and influenza infection [10]. Vaccination status at six of seven sites included plausible self-report rather than medical record documentation, which might result in misclassification of influenza vaccination status for some patients. Second, healthcare seeking behavior has changed during the COVID-19 pandemic, and enrollment of patients with outpatient illness from COVID-19 testing sites might have affected results in uncertain ways. The test-negative design for estimating influenza VE requires validation when multiple vaccine-preventable respiratory viruses are co-circulating. Finally, VE estimates in this report are specific to the prevention of outpatient influenza illness rather than to more severe influenza outcomes (e.g., hospitalization, ICU admission or death), which other study designs may be able to address.

In conclusion, influenza vaccination in 2021–2022 reduced outpatient medically attended acute respiratory illness with cough due to influenza A(H3N2) viruses by approximately one-third overall. Protection afforded by vaccination was comparable to previous A(H3N2)-dominant seasons before the COVID-19 pandemic [13]. To provide better antigenic match to the A(H3N2) viruses that circulated during the 2021–2022 season, the A(H3N2) component for 2022–2023 Northern Hemisphere influenza vaccines was updated to include reference viruses representing the 3C.2a1b.2a.2 subclade. If subclade 2a.2-like viruses continue to circulate, the updated A(H3N2) vaccine component representing the 3C.2a1b.2a.2 subclade may provide improved protection among subclade 2a.2-like viruses during the 2022–2023 influenza season.

## Data Availability

All data produced in the present study are available upon reasonable request to the authors

